# Can we predict early 7-day readmissions using a standard 30-day hospital readmission risk prediction model?

**DOI:** 10.1101/19012468

**Authors:** Sameh N. Saleh, Anil N. Makam, Ethan A. Halm, Oanh Kieu Nguyen

## Abstract

Despite focus on preventing 30-day readmissions, early readmissions (within 7 days of discharge) may be more preventable than later readmissions (8-30 days). We assessed how well a previously validated 30-day readmission prediction model predicts 7-day readmissions. We re-derived model coefficients for the same predictors as in the original 30-day model to optimize prediction of 7-day readmissions. We compared model performance and compared differences in strength of model factors between the 7-day model to the 30-day model. While there was no substantial change in model performance between the original 30-day and the re-derived 7-day model, there was significant change in strength of predictors. Characteristics at discharge were more predictive of 7-day readmissions, while baseline characteristics were less predictive. Improvements in predicting early 7-day readmissions will likely require new risk factors proximal to the day of discharge.

## Background

Despite intense focus on preventing 30-day readmissions, early readmissions within the first 7 days of hospital discharge may be more preventable than later readmissions (8-30 days post-discharge) [1–6]. Early readmissions are more closely related to potential gaps in care during the index hospitalization [4] or reflect premature discharge. Identifying patients at risk for early, rather than later readmissions may be a more effective strategy to tailor resource-intensive transitional care interventions to prevent readmissions. However, current risk prediction models only identify patients at risk for 30-day readmission and often fail to use electronic health record (EHR) data to allow for real-time operationalization of the model. Therefore, we assessed how well a validated 30-day EHR-based readmission risk prediction model [7] would predict early 7-day readmissions, and whether there were differences in the strength of predictors for 7-day versus 30-day readmissions.

## Methods

We conducted an observational cohort study of consecutive hospitalizations by adults ≥18 years from November 2009 to October 2010 using electronic health record (EHR) data from 6 diverse hospitals in north Texas, including safety-net, academic, and community sites. Details of this cohort have been previously published [7]. The primary outcome was all-cause non-elective 7-day hospital readmissions within a 100-mile radius of Dallas, Texas. We split our cohort 50-50 into derivation and validation sets. We used the derivation cohort to re-derive model coefficients for the same predictors from our previously validated 30-day readmission model (also developed from the same cohort) to optimize prediction of 7-day readmissions [7]. We used the validation cohort to compare the discrimination (C-statistic) and calibration of our 7-day readmission model with our original 30-day model to predict 7-day readmissions. We calculated the categorical net reclassification improvement (NRI), which is the absolute net gain in correctly reclassified predictions of high (top 2 risk quintiles) and low risk (bottom 3 quintiles) for the 7-day readmission model compared to the 30-day model [8]. To examine which factors were more (or less) predictive of 7-day readmissions, we evaluated the percent change in coefficients between the two models, using the 30-day model as reference.

## Results

Of 32,922 index hospitalizations among unique patients, 4.4% had a 7-day readmission and 12.7% had a 30-day readmission. Patients with a 7-day readmission tended to be older (65 vs. 62 years old, p≤0.001), have Medicaid (10.4% vs 6.5%, p≤0.001), more prior ED visits (mean 1.3 vs. 0.6 visits in last 12 months, p≤0.001), greater comorbidity burden (mean Charlson comorbidity score of 1.7 vs. 0.9, p≤0.001), and longer length of stay (median 5 vs. 4 days, p≤0.001). Our original 30-day model had modestly lower discrimination for predicting 7-day versus 30-day readmission (C-statistic of 0.66 vs. 0.69, p≤0.001). Our 7-day readmission model had similar discrimination as the original 30-day model for predicting 7-day readmissions (C-statistic of 0.66, p=0.38) but improved calibration, particularly for the highest risk quintile (**Figure 1B**). The 7-day model did not have better reclassification (NRI=0.006, 95% CI: −0.104 – 0.116).

**Figure 1.**
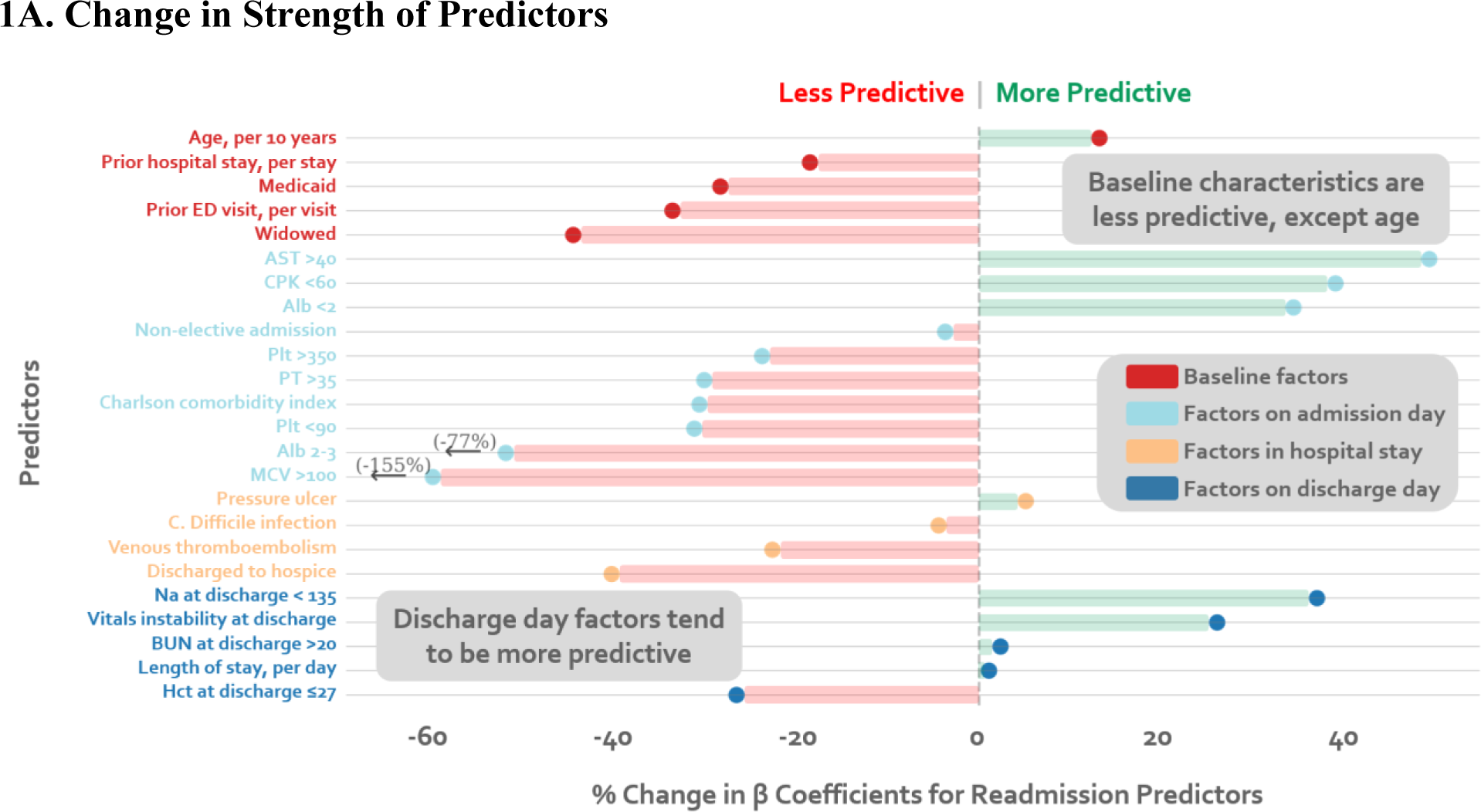
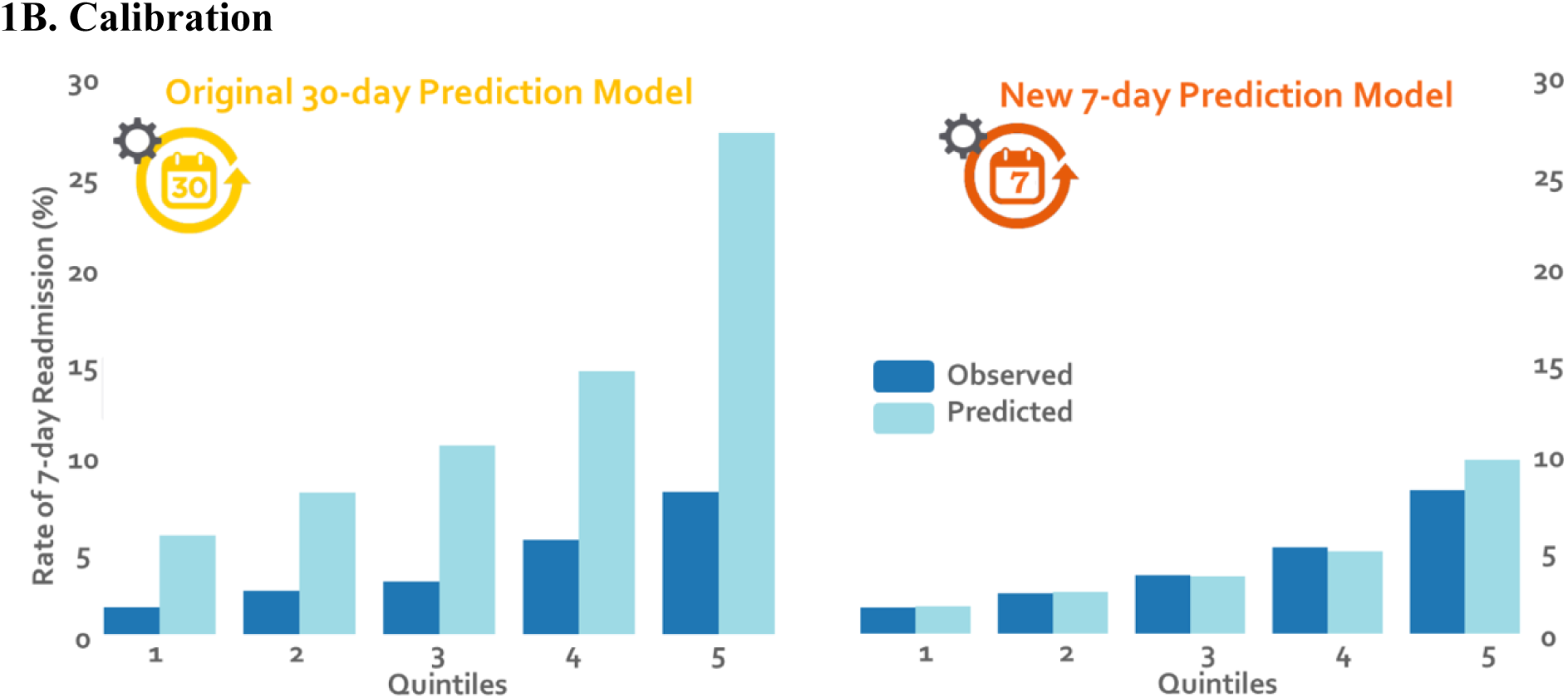
Model Performance of the 7-day versus 30-day Readmission Models. The percent change in β coefficients between the original 30-day model and the re-derived 7-day model is shown for each predictor included in the model. Predictors are grouped according to the timing of their availability, including baseline characteristics prior to the index hospitalization (red dot), factors on hospital admission (light blue dot), factors during hospital stay (gold dot) and factors on discharge day (dark blue dot). Values to the right of the vertical dashed line at 0, shaded in green, indicate factors that are more predictive of early readmission. Values to the left of the dashed line, shaded in red, indicate factors that are less predictive.

When comparing strength of predictors between the two models, clinical characteristics at discharge, such as sodium and vital sign instability, were more strongly predictive of 7-day readmissions compared to 30-day readmissions. Baseline characteristics (Medicaid, widow, prior utilization), were less predictive of 7-day readmissions. Factors on admission and during the hospital stay also tended to be less predictive (**Figure 1A; Table**).

## Conclusions

A previously validated, multi-condition 30-day EHR-based readmission risk prediction model can also be used to predict 7-day readmissions. Model performance was not substantially different compared to a re-derived 7-day readmission model. Reweighting predictors led to slightly better calibration, but risk stratification and reclassification of risk were similar. While overall model performance was similar, strength of predictors for 7-day versus 30-day readmission differed. Characteristics at discharge were more predictive of early 7-day readmissions, while baseline characteristics were less predictive. This is consistent with prior research suggesting that early readmissions are more likely to be related to clinical stability on discharge than 30-day readmissions [1–6].

Our study benefitted from the large, multicenter diverse cohort and high-quality ascertainment of readmissions beyond the index hospital. The use of rich, ubiquitous EHR data allows for real-time operationalization of the model. Furthermore, since we used the original cohort from which the 30-day readmission model was developed [7], we were uniquely positioned to isolate the ability of a 30-day readmission model to predict early 7-day readmissions by avoiding any differences in model performance stemming from changes in the population itself. Study limitations include uncertain generalizability to other settings and use of data before federal penalties for hospital readmission were in effect.

To further optimize model performance, future 7-day readmission risk prediction models should incorporate additional risk factors proximal to day of discharge such as the quality of transition of care planning (e.g. timely outpatient follow-up, medication reconciliation, and dispensing on discharge). Further optimizing risk prediction would enable hospitals to more efficiently target and reduce those readmissions that are potentially the most preventable.

**Table 1.** Comparing Strength of Predictors of 30-day vs. 7-Day Readmissions ^a^

|  | Adjusted Odds Ratio (95% CI) |  |
| --- | --- | --- |
|  | Original 30-day<br>Readmission Model | New 7-day<br>Readmission Model |
| **Baseline factors** |  |  |
| *Demographic characteristics* |  |  |
| Age in years, per 10 years | 1.07 (1.04 - 1.10) | 1.08 (1.03 - 1.14) |
| Widow | 1.27 (1.11 - 1.45) | 1.13 (0.92 - 1.40) |
| Medicaid | 1.55 (1.31 - 1.83) | 1.37 (1.06 - 1.78) |
| *Utilization History* |  |  |
| Prior ED visit, per visit | 1.04 (1.02-1.06) | 1.03 (1.01 - 1.04) |
| Prior hospitalization, per hospitalization | 1.16 (1.12-1.20) | 1.13 (1.08 - 1.18) |
| **Factors from first day of hospitalization** |  |  |
| Nonelective admission | 1.40 (1.09 - 1.80) | 1.42 (1.22 - 1.65) |
| Charlson Comorbidity Index, per point | 1.04 (1.01 - 1.08) | 1.06 (1.04-1.09) |
| *Laboratory abnormalities within 24 hours of admission* |  |  |
| Albumin <2 g/dL | 1.52 (1.05 - 2.21) | 1.75 (1.06 - 2.87) |
| Albumin 2-3 g/dL | 1.20 (1.06 - 1.36) | 1.04 (0.86 - 1.27) |
| Aspartate aminotransferase >40 U/L | 1.21 (1.06 - 1.38) | 1.34 (1.09 - 1.63) |
| Creatine phosphokinase <60 mcg/L | 1.28 (1.11 - 1.46) | 1.40 (1.14 -1.72) |
| Mean corpuscular volume >100 fL/red cell | 1.32 (1.07 - 1.62) | 0.86 (0.60 - 1.23) |
| Platelets <90 x 10<sup>3</sup>/μL | 1.56 (1.23 - 1.97) | 1.36 (0.94 - 1.96) |
| Platelets >350 x 10<sup>3</sup>/μL | 1.24 (1.08 - 1.44) | 1.18 (0.94 - 1.49) |
| Prothrombin time >35 seconds | 1.92 (1.73 -2.90) | 1.57 (0.84 -2.94) |
| **Factors from hospital stay** |  |  |
| Discharge to hospice | 0.23 (0.45 - 1.85) | 0.41 (0.20 - 0.86) |
| *Hospital complications* |  |  |
| Clostridium difficile infection | 2.03 (1.18 - 3.48) | 1.96 (0.96 - 4.00) |
| Pressure ulcer | 1.64 (1.15 - 2.34) | 1.68 (1.01 - 2.79) |
| Venous thromboembolism | 1.55 (1.03 - 2.32) | 1.40 (0.76 - 2.58) |
| **Factors on discharge day** |  |  |
| *Laboratory abnormalities at discharge* |  |  |
| Blood urea nitrogen >20 mg/dL | 1.37 (1.24 - 1.52) | 1.38 (1.17 - 1.62) |
| Sodium <135 mEq/L | 1.34 (1.18 - 1.51) | 1.49 (1.24 - 1.79) |
| Hematocrit ≤ 27% | 1.22 (1.05 - 1.41) | 1.16 (0.92 - 1.46) |
| Vital sign instability at discharge, per instability | 1.25 (1.15 - 1.36) | 1.32 (1.17 - 1.50) |
| Length of stay, per day | 1.06 (1.04 - 1.07) | 1.06 (1.04 - 1.08) |

Abbreviations: ED = emergency department.
<sup>a</sup> Values reflect adjusted odds ratios and 95% CI for each variable after adjustment for all other variables listed in the table separately for our re-derived early model and our original validated 30-day readmission model. For both models, we included index hospitalizations of patients who were alive 30 days post-discharge. Patients who died in the hospital, were transferred to another hospital, or left against medical advice were excluded.

The new 7-day prediction model had better calibration than the original 30-day prediction model across all quintiles of risk, but risk stratification was similar.

## Data Availability

The data that support the findings of this study are available from UT Southwestern but restrictions apply to the availability of these data, which were used under approval for the current study, and so are not publicly available. Data are however available from the authors upon reasonable request and with permission of UT Southwestern.

## Contributorship Statement

Study concept and design: SS, ANM, OKN; Data acquisition: EAH; Analysis: SS; Interpretation of data: all authors; Manuscript preparation: SS, ANM, OKN; Critical revision of manuscript and final approval: all authors.

